# Clinical presentation and evolution of COVID-19 in immunosuppressed patients. Preliminary evaluation in a North Italian cohort on calcineurin-inhibitors based therapy

**DOI:** 10.1101/2020.04.26.20080663

**Authors:** Lorenzo Cavagna, Raffaele Bruno, Giovanni Zanframundo, Marilena Gregorini, Elena Seminari, Angela Di Matteo, Teresa Rampino, Carlomaurizio Montecucco, Stefano Pelenghi, Barbara Cattadori, Eleonora Francesca Pattonieri, Patrizio Vitulo, Alessandro Bertani, Gianluca Sambataro, Carlo Vancheri, Valentina Bonetto, Maria Cristina Monti, Elena Ticozzelli, Annalisa Turco, Tiberio Oggionni, Angelo Corsico, Veronica Codullo, Monica Morosini, Massimiliano Gnecchi, Carlo Pellegrini, Federica Meloni

## Abstract

The clinical course of COVID-19 in patients undergoing chronic immunosuppressive therapy is yet poorly known. We performed a monocentric cross-sectional study describing the clinical course of COVID-19 in a cohort of patients from northern Italy treated with calcineurin-inhibitors for organ transplantation or rheumatic diseases. Data were collected by phone call and clinical chart review between March 27^th^- 31^st^ 2020. COVID-19 related symptoms, rynopharingeal swab, therapeutic changes and outcome were assessed in 384 consecutive patients (57% males; median age 61 years, IQR 48-69). 331 patients (86%) received solid organ transplantation (kidney n=140, 36%, heart n=100, 26%, lung n=91, 24%) and 53 (14%) had a rheumatic disease. Calcineurin inhibitors were the only immunosuppressant administered in 46 patients (12%). 14 patients developed a “confirmed COVID-19” (swab positivity) and 14 a “clinical COVID-19” (only typical symptoms). Fever (75%) and diarrhoea (50%) were the most common symptoms. Fourteen patients were hospitalized and 11 have already been dismissed. No patient required start/changes of the O2 therapy or developed superinfection. Only one patient, with metastatic lung cancer, died. In conclusion, COVID-19 showed a mild course in our cohort, with low mortality. Calcineurin inhibitor-based immunosuppressive regimens appear safe in this context and should not be discontinued.

## Introduction

Coronavirus Disease 2019 (COVID-19) has been first reported in China in 2019 and it is related to a new strain of coronavirus, called “Severe Acute Respiratory Syndrome Coronavirus-2” (SARS-CoV-2)(1). Similarly to 2003 SARS-CoV and “Middle-East Respiratory Syndrome” coronavirus (MERS) (2), SARS-CoV-2 severely affects the lungs and COVID-19 patients are at increased risk of developing an acute respiratory distress syndrome (ARDS) which is often fatal (3). In February 2020, the COVID-19 affected Northern Italy regions, determining a high number of hospitalization and intensive care unit admission (4). The two Italian Regions with the highest prevalence are Lombardy and Emilia-Romagna. According to data released by our National Istituto Superiore di Sanità, on 03.31.2020 the incidence of COVID-19 infection in these two regions was higher than 100 symptomatic cases/100.000 inhabitants. At that time point, 54.036 confirmed cases had already been reported in Lombardy and Emilia Romagna. Furthermore, among the reported symptomatic cases, the 21.3% had a severe presentation of the disease and 3% of patients required intensive cares for respiratory impairment. However, testing and/or reporting strategies varied among Italian regions and this might have led to an underestimation of pauci-symptomatic patients and therefore of the real incidence of COVID-19 infection (5). After the outbreak began, the medical community started having concerns about an increased potential risk of infection in patients on immunosuppressive therapy, even if some preliminary case reports showed a mild disease course in this category of patients (6–9). On the other hand, immunosuppressive treatments are being studied as possible therapeutic options in the hyper-inflammatory phase of the COVID-19 (10). Among the immunosuppressants (ISs), calcineurin inhibitors (CNIs) such as cyclosporine (Cys) and tacrolimus (TAC), which are currently used in the setting of transplantation (11) and Rheumatic Disorders (RMDs) (12), are supposed to exert also an antiviral activity. This peculiar characteristic has been evidenced in vitro also on some coronavirus strains (13–16). Given these assumptions, we aimed at evaluating the occurrence of SARS-CoV-2 infection and its clinical manifestations in a cohort of patients referring to our center and currently on CNIs based treatment regimen, for either RMDs or solid organ transplantation.

## Materials and Methods

Between the 27^th^ and the 31^st^ of March 2020, more than one month after the beginning of the COVID-19 outbreak in Italy, we conducted cross-section study on patients undergoing an immunosuppressive treatment including CNIs. Patients referred to the Units of Rheumatology, Respiratory diseases, Nephrology and Cardiac surgery of our Hospital. We selected patients from Northern Italian regions first interested by SARS-CoV-2 infection (Lombardy, Emilia-Romagna, Piedmont and Veneto). The included patients were followed up for RMDs such as antisynthetase syndrome (ASSD), idiopathic inflammatory myopathies (IIMS), systemic lupus erythematosus (SLE), Adult onset Still disease (AOSD) and for Solid Organ transplantation (eg heart, lung and kidney). All patients agreed to participate in this interview and provided the informed consent as approved by our IRB. We carried-out a pre-established phone survey aimed at investigating the occurrence of symptoms and laboratory/radiology findings possibly related to SARS-CoV-2 infection during the 30 days preceding the contact and at identifying patients diagnosed with COVID-19. Data collection for the patients admitted to our Hospital was performed by direct interview and clinical chart review. We assessed the following variables: fever (higher than 37.5 ° C), persistent rhinorrhoea, ageusia, anosmia, headache, nausea, vomiting, diarrhoea, dyspnoea, cough, sore throat, fatigue, arthromyalgias, current treatments, contact with diagnosed COVID-19 cases and, if available, laboratory (C-reactive protein, Lactate dehydrogenase, hypocalcemia, lymphopenia) and chest X-rays results. We retrieved also the most recent CNIs blood basal levels, the time passed from symptoms onset, the occurrence of hospitalizations, with related clinical files, and the final-outcome, defined as: full-recovery, improvement (partial relief from symptoms), not improvement (symptoms unchanged), worsening (onset of new symptoms or worsening of existing ones), death. Data regarding ongoing treatments and changes in the immunosuppressive regimen were also acquired. In patients admitted at our Hospital with SARS-CoV-2 related lung involvement, we assessed serum interleukin-6 (IL-6) by ELISA, high sensitivity C-reactive protein (hs-CRP) by ADVIA Chemistry XPT system and collected all clinical files. Patients were diagnosed with “confirmed COVID-19” in case of rhinopharyngeal swab positivity for SARS-CoV-2. In the absence of rhinopharyngeal swab, we considered a diagnosis of “clinical COVID-19” if: 1) at least 4 typical clinical/laboratory/radiology findings were present; 2) at least 3 typical clinical/laboratory/radiology findings plus a reported contact with a confirmed SARS-CoV-2 patient were present. For both point 1 and 2 it was necessary to rule out other diagnoses explaining the clinical picture. For patients with negative rhinopharyngeal swabs and more than 4 clinical findings of COVID-19, without other possible causes identified, the working group discussed about the classification or not of the patient as a clinical COVID-19, targeting the unanimous agreement on the final diagnosis.

Patients were then considered as having lung involvement in case of bilateral pneumonitis at chest X-rays or new appearance of dyspnoea or its worsening with significant lowering of peripheral oxygen saturation (<92%). We included also a control cohort from Sicily, a non-endemic area for COVID-19 at the time of the survey, composed by patients in follow-up at the ISMETT center of Palermo, and at the University Hospital Policlinico Vittorio-Emanuele of Catania. The aim of the control group was to evaluate whether the occurrence of a clinical picture consistent with our definition of “clinical COVID-19” could be observed in a comparable cohort of patients not exposed to SARS-CoV-2.

## Statistical analysis

Data were reported as absolute numbers and percentages for categorical variables. Numerical variables were described using median and interquartile range. Differences related to quantitative variables between two groups were evaluated by Mann-Whitney U test or unpaired Student t-test as appropriate, while differences related to quantitative variables among more than two groups were evaluated by Kruskall-Wallis test or the Analysis of Variance (ANOVA). Chi-square or Fisher exact tests were applied to explore differential distributions among groups of patients. The occurrence of COVID-19 was expressed as a relative frequency (number of cases at the end of the survey divided by total number of patients), by type of patient and in the whole sample of patients treated with CNIs. A p-value of less than 0.05 was considered as statistically significant. The analyses were performed by one statistician (CM) and one healthcare professional with experience in statistical analysis (LC) using Stata 15 (StataCorp. 2017. Stata Statistical Software: Release 15. College Station, TX: StataCorp LLC.).

## Results

We retrieve data from 384 patients (220 males, 57%) on CNIs based therapy (TAC 214, 56%, Cys 170, 44%), representing the entire cohort of eligible patients from northern Italy followed at our Hospital. Most patients enclosed received a solid organ transplantation (kidney n=140, 36%, heart n=100, 26%, lung n=91, 24%), while 53 patients (14%) suffered from RMDs: 42 patients had ASSD with interstitial lung disease (ILD) (11%), 2 had IIMs (4%), 6 had SLE (12%) and 3 had AOSD (6%). Overall characteristics of the patients enrolled in the study are reported in table 1, while geographic area of referral of the cohort and of the control group is reported in figure 1. All patients followed the national rules about isolation established by our Government since late February 2020 and subsequent updates. Forty-six patients (12%) were treated exclusively with CNIs (TAC n=10, 12%, Cys n=36, 78%) and 338 (88%) also with other ISs in multiple association (corticosteroids n=203, 53%, micophenolate mophetil n=188, 49% everolimus n=65, 17%, azathioprine n=17, 4%, other n=23, 6%). Blood levels of Cys and TAC were found in therapeutic range according to underlying condition for every patient in the previous three months. Unfortunately, it was not possible to re-asses the drugs levels in all patients at the time of the interview. A “confirmed COVID-19” was identified in 14 cases (4%), whereas other 14 patients (4%) were classified as “clinical COVID-19”. We did not observe a gender prevalence in both confirmed (10 males and 4 females, p=0.276) and confirmed + clinical COVID-19 (15 males and 13 females, p=0.679) patients. No substantial differences were observed in COVID-19 frequency between patients in combination therapy vs in CNI alone (confirmed: 3 versus 11 cases, p=0.267; clinical + confirmed: 4 versus 24 cases p=0.696). The symptoms observed in COVID-19 patients are reported in table 2. Symptoms frequency was substantially similar between confirmed and clinical COVID-19, with the exception of diarrhoea, nausea and anosmia, that were more commonly observed in the confirmed COVID-19 cohort (10 vs 1, p<0.001; 8 vs 2, p=0.046; 5 vs 0, p=0.04, respectively). COVID-19 related lung involvement was observed in 11 cases (40% of total COVID-19), the majority (8, 73%) had a “confirmed COVID-19” diagnosis. No patients developed severe respiratory complications, including the 42 (11% of total) RMDs patients with interstitial lung disease (ILD) and the 91 (24% of total) lung transplanted patients.

**Figure 1:**
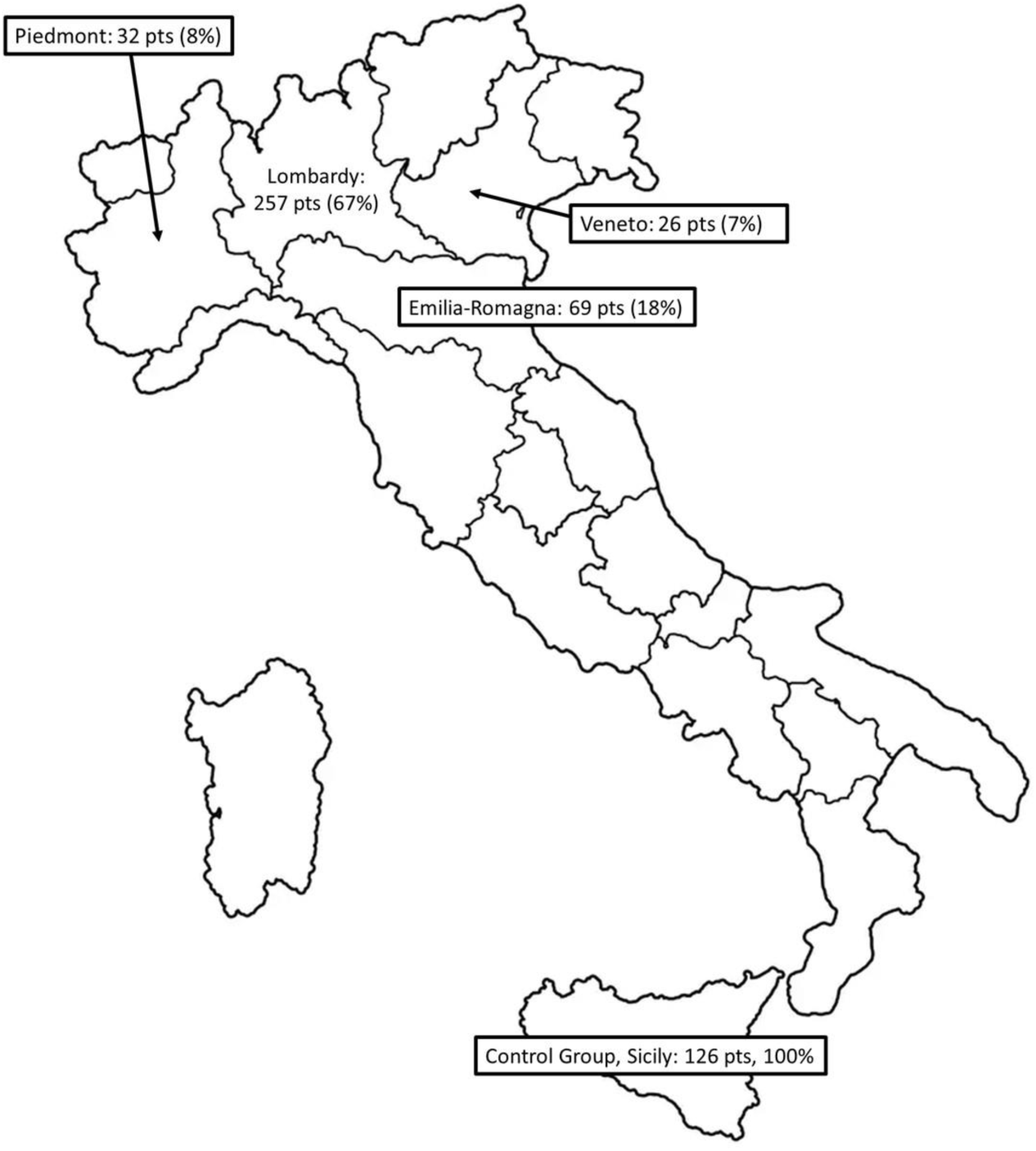
Geographical origin of the patients

**Table 1:** General characteristic of patients evaluated. Legend: COVID-19: Coronavirus Disease 2019; IQR: interquartile range; Tx transplantation; Cys: cyclosporine; Tac: tacrolimus; RMDs: rheumatic diseases; IS: immunosuppressants).

|  | Patients<br>(%) | Median<br>age in<br>years<br>(IQR) | Median<br>follow-up in<br>months<br>(IQR) | Males<br>(% by<br>group) | Cys (%)<br>by<br>group) | Tac (%)<br>by<br>group) | CNIs<br>and<br>other<br>IS (%)<br>by<br>group) | Confirmed<br>diagnosis<br>of COVID-19 (%) | Median<br>age in<br>years<br>(IQR) | Median<br>follow-up in<br>months<br>(IQR) | Males<br>(% by<br>group) | Cys (%)<br>by<br>group) | Tac (%)<br>by<br>group) | CNIs<br>and<br>other<br>IS (%)<br>by<br>group) | Clinical<br>diagnosis<br>of COVID-19 (% of<br>total) | Median<br>age in<br>years<br>(IQR) | Median<br>follow-up in<br>months<br>(IQR) | Males<br>(% by<br>group) | Cys (%)<br>by<br>group) | Tac (%)<br>by<br>group) | CNIs and<br>other IS<br>agents<br>(% of<br>group) |
| --- | --- | --- | --- | --- | --- | --- | --- | --- | --- | --- | --- | --- | --- | --- | --- | --- | --- | --- | --- | --- | --- |
| **RMDs** | 53 (14) | 61 (48-69) | 41 (17-81) | 15 (28) | 53<br>(100) | 0 (0) | 30 (57) | 0 (0) | - | - | - | - | - | - | 6 (43) | 57 (48-59) | 99 (17-140) | 2 (33) | 6 (100) | 0 (0) | 5 (40) |
| **Lung Tx** | 91 (24) | 58 (48-64) | 116 (46-187) | 54 (59) | 11 (12) | 80 (88) | 91<br>(100) | 3 (21) | 65 (62-75) | 60 (1-266) | 2 (67) | 1 (33) | 2 (67) | 3 (100) | 4 (29) | 57<br>(47.5-67) | 143.5<br>(84.5-232) | 2 (50) | 0 (0) | 4 (100) | 4 (30) |
| **Heart**<br>**Tx** | 100 (26) | 63 (53-72.5) | 144 (72-228) | 61 (61) | 63 (63) | 37 (37) | 78 (78) | 5 (36) | 63 (53-67) | 60 (48-144) | 3 (60) | 2 (40) | 3 (21) | 3 (60) | 2 (14) | 62.5<br>(57-68) | 72 (60-84) | 0 (0) | 2 (100) | 0 (0) | 2 (15) |
| **Kidney**<br>**Tx** | 140 (36) | 55 (48-62.5) | 62.5 (32.5-116.5) | 90 (64) | 43 (31) | 97 (69) | 139<br>(99) | 6 (43) | 57.5 (51-64) | 95 (46-159) | 5 (83) | 3 (50) | 3 (21) | 5 (83) | 2 (14) | 38 (35-41) | 37 (17-57) | 1 (50) | 0 (0) | 2 (100) | 2 (15) |
| **Overall** | 384<br>(100)* | 58 (49-66) | 83.5 (39-156) | 220<br>(57) | 170<br>(44) | 214<br>(56) | 338<br>(88) | 14 (100) | 60.5 (46-159) | 62.5<br>(53-67) | 10 (71) | 6 (43) | 8 (57) | 11 (79) | 14 (100)* | 84 (40-140) | 87.5 (57-140) | 6 (43) | 9 (64) | 6 (43) | 13 (94) |

**Table 2:** Symptoms prevalence in the COVID-19 cohort. (total COVID-130=28, confirmed COVID-19=14, clinical COVID-19=14). * Chi2 test; other Fisher exact test

| Symptoms | Number of all COVID19 patients (%) | Number of confirmed-COVID19 patients (%) | clinical-COVID19 (% on 14 patients) | P |
| --- | --- | --- | --- | --- |
| Fever | 21 (75) | 12 (86) | 9 (64) | 0.190* |
| Fatigue | 16 (57) | 10 (71) | 6 (43) | 0.127* |
| Cough | 14 (50) | 9 (64) | 5 (36) | 0.257 |
| Rhinorrea | 14 (50) | 5 (35) | 9 (64) | 0.257 |
| Headache | 13 (46) | 8 (57%) | 5 (36) | 0.449 |
| Diarrhoea | 11 (39) | 10 (71) | 1 (7) | 0.001 |
| Nausea | 10 (36) | 8 (57%) | 2 (14) | 0.046 |
| Sore throat | 6 (21) | 4 (28) | 2 (14) | 0.648 |
| Ageusia | 6 (21) | 4 (28) | 2 (14) | 0.648 |
| Arthromyalgias | 11(39) | 4 (18) | 7 (50) | 0.440 |
| Dysphnea | 5 (18) | 3 (21) | 2 (14) | 1.000 |
| Anosmia | 5 (18) | 5 (36) | 0 (0) | 0.041 |
| Conjunctivitis | 4 (14) | 2 (14) | 2 (14) | 1.000 |

The median symptom duration in the whole COVID-19 cohort was 14 days (IQR 10-19.5). Symptoms duration accordingly to COVID-19 status is reported in table 3 and 4. A large number of patients improved (n=19, 68%). We registered only one case of death (4% of confirmed + clinical COVID-19 group) in a patient also suffering from lung metastatic cancer. The remaining 8 patients remained stable. The clinical characteristics and outcome of confirmed + clinical COVID-19 have been reported respectively in table 3 and table 4, respectively. Fourteen transplanted patients (4% of total, 12 confirmed COVID-19 and 2 clinical COVID-19) have been admitted to our Hospital, mainly for fever and diarrhoea and 11 (79% of hospitalized patients) have already been discharged after recovering from the disease. At the admission, CNIs were maintained, whit a tapering of other ongoing ISs in all cases. After ISs reduction no transplanted patients developed signs or symptoms of acute rejection. Conversely, no treatment changes were made in non-hospitalized patients. Baseline IL-6 serum levels (normal value 0-3.12 pg/ml) in patients admitted to our Hospital were in median 41 pg/ml (IQR 20.4 41.1), whereas median hs-CRP (upper normal limit: 0.5 pg/ml) was 5.6 mg/dl (IQR 3.3-10.1). Interestingly, IL-6 levels were significantly lower compared with a group of COVID-19 patients (median 235.5, IQR 163.1-235.5; p<0.005), matched for sex, age, and time from symptoms onset, while hs-CRP was comparable (median 6.1, IQR 3.0-14.6; p=0.399). Of note, these patients were admitted mainly for respiratory impairment and they were subsequently treated with tocilizumab for disease progression. All admitted patients started HCQ and azithromycin (17), as for internal protocol at that specific time period. No superimposed infections have been reported in hospitalized patients. None of the patients required initiation or modification (when already administered at home) of high flow O2 therapy or developed ARDS.

**Table 3:** Characteristic of the 14 patients with confirmed COVID-19. *the patient died for lung metastatic cancer. Legend: COVID-19: Coronavirus Disease 2019; RMDs: Rheumatic diseases; Tx transplantation; ARDS: acute respiratory distress syndrome

| Condition (number, % of overall) | Median time from symptoms onset in days (IQR) | Isolation at home (% of the group) | Hospitalization (% of the group) | Clinical/radiological lung involvement (% of the group) | ARDS (% of the group) | Stable (% of the group) | Improved (% of the group) | Worsened (% of the group) | Death (% of the group) |
| --- | --- | --- | --- | --- | --- | --- | --- | --- | --- |
| **RMDs (0, 0)** | - | - | - | - | - | - | - | - | - |
| **Lung (3, 21)** | 15 (10-18) | 1 (33) | 2 (67) | 2 (67) | 0 (0) | 0 (0) | 3 (100) | 0 (0) | 0 (0) |
| **Heart (5, 36)** | 12 (10-15) | 0 (0) | 5 (100) | 1 (20) | 0 (0) | 4 (67) | 0 (0) | 0 (0) | 1 (33)* |
| **Kidney (6, 43)** | 12.5 (10-15) | 1 (20) | 5 (80) | 5 (83) | 0 (0) | 3 (50) | 3 (50) | 0 (0) | 0 (0) |
| **Overall (14, 100)** | 13.5 (10-15) | 2 (14) | 12 (86) | 8 (57) | 0 (0) | 7 (50) | 6 (43) | 0 | 1 (7) |

**Table 4:** Characteristic of the 14 patients with clinical COVID-19. Legend: COVID-19: Coronavirus Disease 2019; RMDs: rheumatic diseases; Tx transplantation; ARDS: acute respiratory distress syndrome

| **Disease Tx (number,**<br>**% of overall)** | **Median time from**<br>**symptoms onset in**<br>**days (IQR)** | **Isolation at home (%**<br>**of the group)** | **Hospitalization (%**<br>**of the group)** | **Clinical/radiological**<br>**lung involvement (% of**<br>**the group)** | **ARDS (% of**<br>**the group)** | **Stable (%**<br>**of the**<br>**group)** | **Improved**<br>**(% of the**<br>**group)** | **Worsened**<br>**(% of the**<br>**group)** | **Death (% of**<br>**the group)** |
| --- | --- | --- | --- | --- | --- | --- | --- | --- | --- |
| **RMDs (6, 44)** | 20 (19-21) | 6 (100) | 0 (0) | 1 (17) | 0 (0) | 0 (0) | 6 (100) | 0 (0) | 0 (0) |
| **Lung (4, 28)** | 15 (10-20) | 3 (75) | 1 (25) | 1 (25) | 0 (0) | 0 (0) | 4 (100) | 0 (0) | 0 (0) |
| **Heart (2, 14)** | 8 (7-9) | 2 (100) | 0 (0) | 0 (0) | 0 (0) | 1 (50) | 1 (50) | 0 (0) | 0 (0) |
| **Kidney (2, 14)** | 8 (5-11) | 1 (50) | 1 (50) | 1 (50) | 0 (0) | 0 (0) | 2 (100) | 0 (0) | 0 (0) |
| **Overall (14, 100)** | 15 (9.5-20) | 12 (86) | 2 (14) | 3 (21) | 0 (0) | 1 (7) | 13 (93) | 0 (0) | 0 (0) |

## Control group

The control group included 126 patients (67 males, 53%, p=0.419 versus our cohort), of which 111 (88%) were lung transplanted and 15 had RMDs complicated by ILD (12%). Overall median age of the control group was 55 years (IQR 41-65), that was not statistically different from what observed in our cohort (p=0.125). Our criteria of “clinical COVID-19” were not satisfied by any of the patient of the control group. During the period of observation, none of these patients was hospitalized or reported new appearance or worsening of dyspnea.

## Discussion

The effect of SARS-CoV-2 infection in patients on immunosuppressive treatment is still matter of debate. In our study we observed that the majority of our patients strictly followed the steadily increasing rules on personal isolation suggested by our government and reinforced by referring clinicians. Although the frequency of confirmed + clinical COVID-19 was quite high in our cohort, the clinical presentation was generally mild. Even patients with lung involvement did not progress to ARDS. One possible explanation for this increased frequency could be the tight control routinely applied to this cohort, that could facilitate the diagnosis, possibly at an early stage of the disease. Furthermore, as already mentioned, the true frequency of COVID-19 in the general population of Italy is still not well established and in some areas it could be similar to that we reported (5).

Another point to highlight is that the short-time outcome of our patients was favourable. Indeed, the clinical condition of a large number of patients improved and no one reported worsening of the symptoms. Only one patient had a negative outcome, but it was considered only partially attributable to SARS-CoV-2 infection. Despite the short observation time (14 days), our data on favourable outcome should be considered relevant, since, according to a recent report of the Istituto Superiore di Sanità (https://www.epicentro.iss.it/coronavirus/bollettino/Report-COVID-2019_26_marzo_eng.pdf), median time between symptoms onset and fatality is of 9 days. Interestingly, no patients developed superimposed infections despite immunosuppression. We observed confirmed COVID-19 cases only in the setting of transplantation and not in RMDs patients. In the RMDs cohort, despite the occurrence of an underlying ILD in the majority of cases, the only patients developing transient respiratory symptoms had a diagnosis of SLE without a previous lung involvement. Two hospitalized patients had negative rhinopharyngeal swabs, without further attempt of diagnosis by analysing other biological samples. Since other possible diagnoses were ruled out, after collegial discussion, we included these patients in the clinical COVID-19 group. The surprisingly low severity of COVID-19 infection in our cohort might have three possible explanations: 1) as previously reported, the tight control we routinely adopted on this cohort may have facilitated not only the early diagnosis, but also the early hospitalization and treatment of the patients, 2) it is possible that the long term immunosuppressive regimen used in our cohort might have influenced the clinical course of the disease by preventing the occurrence of the huge alveolar macrophage activation with consequent release of pro-inflammatory cytokines that has been described in the context of SARS-COVID-19 (10). In fact, although hs-CRP levels were similar between confirmed COVID-19 with and without immunosuppression, IL-6 levels were much lower in the immunosuppressed group, in line with this hypothesis. 3) we cannot exclude a direct antiviral activity of some of the ISs used. Both Cys and TAC inhibit viral replication in a number of strains of CoV, including SARS-CoV, through the inhibition of peptidyl-prolyl cis-trans isomerases, such as cyclophilin A and FK506-binding proteins, that are cellular interaction partners of CoV non-structural protein 1 (Nsp1) (13–15). If we hypothesize that Cys and TAC exert antiviral activity also towards SARS-CoV-2, we may suppose an added value of CNIs with respect to pure ISs, because theoretically able to reduce the viral load or the risk of viral load increase. On this basis, it is reasonable to add also CNIs in the list of ISs possible therapeutic options for COVID-19 (18).

Our study has some limitations. First, a short temporal window was considered, limiting the number of possible events recorded. Indeed, we cannot exclude that some of the contacted asymptomatic patients could develop the disease in the next days or weeks or that some of the patients could worsen or die subsequently. Second, we assessed most patients only by a phone-call, without a direct visit due to the established quarantine and only a few patients had recent blood tests available, which probably resulted in missing some asymptomatic cases. Third, the definition of clinical COVID19 may be matter of discussion. However, the substantial lack of patients with similar characteristic in the control group is surely a factor that strengthen our choice to add this class of patients in the analysis. Fourth, the control group was not completely comparable to the study group, since it included only patients with RMDs or lung transplantation. Nevertheless, we believe that this limit doesn’t really affect our results, since treatments regimen are comparable for all transplanted patients. Fifth, we could not find a completely equal control group of not immunosuppressed patients for both confirmed and clinical COVID-19 and for both outpatients and inpatients. Indeed, during the period of our observation, no immunosuppressed COVID-19 patients had generally been admitted to our Hospital with a more compromised respiratory picture compared to our cohort. The reason is that all admitted patients belonging to our cohort were solid organ transplanted and admitted mainly for fever and diarrhoea, and not for respiratory symptoms.

This preliminary report suggests that COVID-19 course in patients treated with CNIs alone or in association with other ISs appears to be generally mild, also in the case of SARS-CoV-2 related lung involvement or of previously diagnosed ILD. Furthermore, it is important to underline that no patient developed infectious complication. Substantially, CNIs based chronic immunosuppressive regimens do not increase the risk of a more severe course of COVID-19 or complications. Our result indicates the need of a careful discussion on the effect of immunosuppression on COVID-19. The substantial lack of ARDS in our cohort may also suggest a protective action of immunosuppression in these patients, at least for one of the most dreadful complications of the disease. This might be especially true in the case of CNIs, which, besides their action on immune system, could also exert an antiviral action on coronavirus.

## Data Availability

Data available on request due to privacy/ethical restrictions

